# An End-to-End Synthetic Oncology Clinical Trial Framework Integrating Radiographic Response, Circulating Tumor DNA, Safety, and Survival for Decision-Oriented Clinical Data Science

**DOI:** 10.64898/2026.04.07.26350297

**Authors:** Mark I.R. Petalcorin

## Abstract

**Background:** Modern oncology development depends on integrating radiographic response, molecular biomarkers, treatment exposure, safety, and survival endpoints, yet access to well-structured patient-level trial data is often limited.

**Methods:** We developed a synthetic, literature-informed phase II randomized oncology trial framework that followed the sequence Patient → Data → Dataset → Analysis → Tables/Figures → Decision. A cohort of randomized patients was simulated with baseline demographic and disease features, longitudinal tumor measurements, circulating tumor DNA, inflammatory and exploratory biomarkers, adverse events, treatment exposure, and survival outcomes. Raw source datasets were transformed into SDTM-like domains and ADaM-like analysis datasets, then analyzed for baseline characteristics, exposure, best overall response, survival, subgroup hazard ratios, longitudinal tumor and biomarker changes, exposure-response, and safety.

**Results:** The treatment arm showed a coherent efficacy signal across multiple analytical layers. Treatment increased objective response and clinical benefit, reduced tumor burden over time, and prolonged survival. Median overall survival increased from 135 days in the control arm to 288 days in the treatment arm, with an approximate hazard ratio of 0.661 (95% CI, 0.480-0.911; p = 0.011). Median progression-free survival increased from 116 to 208 days, with an approximate hazard ratio of 0.601 (95% CI, 0.418-0.864; p = 0.006). Circulating tumor DNA showed a more favorable trajectory in treated patients and aligned directionally with radiographic and survival benefit. Safety analyses showed increased treatment-related toxicity, but the overall safety profile remained interpretable and compatible with continued development.

**Conclusions:** This study demonstrates that a synthetic, literature-informed oncology trial can reproduce a biologically plausible and analytically coherent efficacy-safety signal architecture across radiographic, molecular, and time-to-event endpoints, providing a decision-oriented prototype for translational oncology clinical data science.

## Introduction

Cancer is a dynamic disease state in which altered cell proliferation, impaired death signaling, stress adaptation, immune evasion, metabolic rewiring, angiogenesis, and genomic instability become integrated into a self-sustaining biological system (Hanahan & Weinberg, 2000; Hanahan, 2022). At the molecular level, malignant cells accumulate genetic and epigenetic changes that reprogram signaling pathways controlling cell-cycle progression, apoptosis, DNA repair, nutrient acquisition, redox balance, and tissue invasion. At the systems level, those cell-intrinsic changes reshape the tumor microenvironment, distort vascular supply, alter stromal and immune-cell behavior, and ultimately perturb whole-body physiology through inflammation, cachexia, organ dysfunction, and metabolic stress (Hanahan, 2022). In practical oncology, this means that the biologic meaning of treatment response cannot be fully captured by a single variable. Tumor size, blood biomarkers, adverse events, and survival endpoints each measure a different layer of the same evolving disease process.

For this reason, contemporary cancer drug development evaluates efficacy and safety through an integrated framework that combines radiographic response, time-to-event endpoints, treatment exposure, laboratory monitoring, and increasingly, serial molecular biomarkers. In solid tumors, RECIST 1.1 remains the dominant imaging framework for standardized assessment of tumor burden and objective response, because changes in measurable lesions provide a practical and reproducible representation of antitumor activity at the organism level (Eisenhauer et al., 2009). Yet RECIST has recognized limitations. It compresses a heterogeneous biological process into size-based thresholds and may miss early molecular response, immune-related atypical patterns, or biologically important residual disease that is not yet visible on imaging (Ko et al., 2021). Likewise, endpoints such as objective response rate, progression-free survival, and overall survival capture different aspects of therapeutic effect and often diverge when a treatment shrinks tumors without changing long-term disease ecology, or delays progression without extending life substantially (Nie et al., 2019). These realities have driven increasing interest in analytical strategies that connect patient-level variables across time and across biological scales rather than treating each endpoint as a separate silo.

Among emerging biomarkers, circulating tumor DNA, ctDNA, is especially important because it provides a minimally invasive molecular readout of tumor burden, clone persistence, and treatment response. ctDNA consists of tumor-derived DNA fragments shed into the circulation through apoptosis, necrosis, and other forms of tumor-cell turnover. Because it reflects tumor-derived nucleic acids in real time, ctDNA can capture biological change earlier than conventional radiographic endpoints in some settings and can sample clonal heterogeneity more broadly than a single tissue biopsy (Bittla et al., 2023; Thompson et al., 2023). Recent studies and meta-analyses have shown that higher ctDNA burden is associated with poorer prognosis and that early ctDNA dynamics can correlate with radiographic response and long-term outcome in solid tumors, including advanced disease settings (Assaf et al., 2023; Kansara et al., 2023; Li et al., 2023). Thus, ctDNA is not merely a supplementary biomarker but also a candidate mechanistic bridge linking subclinical molecular response to later imaging and survival outcomes. A flagship oncology data science workflow benefits from including both categorical biomarker stratification and continuous ctDNA trajectories because together they emulate the translational logic of companion diagnostics and response monitoring.

Cancer progression is also governed by host-level systemic biology, not only by lesion-centric growth. Biomarkers such as lactate dehydrogenase, C-reactive protein, and albumin are clinically useful because they reflect how malignant growth interacts with metabolism, inflammation, tissue injury, and physiologic reserve. LDH is closely tied to glycolytic metabolism and hypoxia-associated stress, and elevated circulating LDH has shown consistent adverse prognostic value across many solid tumors (Petrelli et al., 2015). CRP reflects systemic inflammatory signaling, while albumin decreases in the setting of sustained inflammation, altered hepatic protein synthesis, nutritional decline, and advanced disease burden. Inflammation-based prognostic frameworks, including CRP- and albumin-centered scores, repeatedly stratify survival across cancer types because they summarize a biologically important interface between tumor behavior and host response (McMillan, 2013; Shibutani et al., 2016). At the molecular level, these markers reflect cytokine-driven acute-phase responses, catabolic stress, metabolic reprogramming, and tissue damage. At the systems level, they indicate that cancer behaves as a whole-body disorder whose clinical trajectory cannot be inferred from tumor measurements alone.

One of the most important molecular nodes linking local tumor biology to systemic deterioration is interleukin-6, IL-6. IL-6 is a pleiotropic cytokine that can activate JAK/STAT3-centered signaling programs involved in proliferation, survival, invasion, angiogenesis, stem-like behavior, immune evasion, and therapeutic resistance (Guo et al., 2012; Kumari et al., 2016). Persistent IL-6 activity can therefore sustain tumor-supportive ecosystems even in the presence of transient radiographic response. This has practical implications for clinical trial interpretation. A therapy may induce short-term lesion shrinkage while failing to durably suppress the inflammatory and adaptive circuits that drive recurrence, progression, and clinical decline. For this reason, longitudinal biomarker panels that include inflammatory mediators can provide biologically meaningful context for survival and response analyses. In a systems-oriented oncology model, IL-6 is not simply an inflammatory marker, it is a signaling hub that links tumor cells, stromal cells, immune cells, vascular remodeling, and organism-level sickness behavior.

Durable cancer control depends not only on direct cytotoxic or cytostatic effects on tumor cells, but also on how therapy reshapes the tumor microenvironment. The cancer-immunity cycle provides a useful framework for understanding this interaction: tumors are controlled when cancer antigens are released, presented, recognized, and converted into effective T-cell-mediated killing, and when suppressive signals within the microenvironment do not abort that sequence (Chen & Mellman, 2013; Mellman et al., 2023). PD-L1 has become a widely used, though imperfect, biomarker within this ecosystem because it reflects one dimension of immune context and can correlate with response to immune-checkpoint inhibition in selected settings (Doroshow et al., 2021). VEGF-dependent angiogenic biology also plays a central role, because abnormal vasculature alters oxygen delivery, nutrient flux, immune-cell trafficking, and drug penetration while supporting tumor persistence and escape (Tugues et al., 2011; Vasudev & Reynolds, 2014). Consequently, biomarkers related to immune activation, immune suppression, and angiogenesis can help interpret why some patients convert early tumor shrinkage into prolonged survival whereas others progress despite apparent initial activity.

These biological realities motivate an end-to-end analytical architecture in oncology in which patient-level baseline factors, longitudinal measurements, standardized datasets, and derived analysis datasets are explicitly connected to interpretable efficacy and safety outputs. In such a framework, the patient is the starting point, raw clinical and biomarker observations become structured data, structured data become analysis-ready datasets, and those datasets support tables, figures, and decisions. This workflow reflects how real development programs operate, particularly in early- and mid-phase oncology trials where the objective is not only to estimate efficacy but also to determine whether a therapeutic signal is coherent enough to justify escalation into a larger confirmatory or biomarker-enriched study. Synthetic, literature-informed datasets are useful in this context because they allow investigators to prototype these integrated workflows without accessing confidential patient records. When carefully constructed, such datasets can reproduce the signal architecture of real clinical development, including baseline heterogeneity, longitudinal response, biomarker dynamics, exposure variability, safety trade-offs, and time-to-event outcomes.

Accordingly, the present work was designed as a synthetic, literature-informed phase II oncology study that links baseline clinical risk, radiographic tumor response, ctDNA dynamics, inflammatory and exploratory biomarkers, treatment exposure, adverse events, and survival outcomes within a single decision-oriented analysis pipeline. The objective was not to claim clinical efficacy for any real investigational therapy, but to create a biologically plausible and analytically rigorous framework that mirrors modern oncology development. By integrating patient-level baseline features with longitudinal tumor and biomarker data, the study aims to demonstrate how coherent evidence can emerge across radiographic, molecular, safety, and time-to-event layers. In this way, the project functions as both a computational simulation and a translational data science framework, converting the sequence Patient → Data → Dataset→ Analysis → Tables/Figures → Decision into a practical workflow that captures the molecular and systems-level complexity of cancer therapy.

## Methods

### Study design and overall analytical framework

This study used a synthetic, literature-informed phase II randomized oncology trial framework designed to emulate a modern solid-tumor development program while avoiding the use of real patient records. The analytical architecture followed the sequence Patient → Data → Dataset → Analysis → Tables/Figures → Decision, with the goal of generating a biologically plausible and decision-oriented clinical data science workflow. The study was constructed to mirror the logic of oncology trials in which baseline clinical risk, longitudinal tumor assessments, circulating biomarkers, treatment exposure, safety events, and time-to-event outcomes are integrated into standardized and analysis-ready datasets for downstream efficacy and safety evaluation (Eisenhauer et al., 2009; Assaf et al., 2023).

The synthetic trial was specified as a two-arm randomized study with 1:1 allocation to control or treatment, with follow-up extending to approximately 24 months. The core endpoints were overall survival, progression-free survival, best overall response, objective response rate, grade 3 or higher adverse events, biomarker change over time, and tumor burden change over time. The intent was not to recreate any single published trial exactly, but to reproduce a realistic signal structure benchmarked to commonly observed oncology trial patterns reported in peer-reviewed clinical and translational literature.

### Synthetic patient population and baseline variable generation

Each patient contributed one baseline record to the source demographics dataset. Synthetic patients were assigned unique identifiers and site identifiers, with study participation distributed across multiple international sites. Baseline variables included country, sex, age, race, ethnicity, anthropometric measures, ECOG performance status, smoking status, biomarker status, baseline lactate dehydrogenase, C-reactive protein, albumin, circulating tumor DNA fraction, and baseline tumor burden.

Continuous baseline variables were simulated from truncated normal or log-normal distributions selected to reflect clinically plausible oncology cohorts. Age was centered in the early 60s, with truncation to adult oncology ranges. Height and weight distributions were sex-specific, and body mass index was derived from weight and height. ECOG performance status was sampled as an ordinal baseline functional variable. Biomarker status was simulated as a binary positive or negative subgrouping variable to support companion diagnostic-style analyses. Baseline LDH and CRP were simulated from right-skewed distributions to reflect the real-world tendency of inflammatory and tumor-burden-linked biomarkers to show long upper tails. Baseline albumin was simulated from a truncated normal distribution centered near the lower end of the healthy adult range, reflecting the mild systemic compromise often present in oncology patients. Baseline ctDNA fraction was generated as a bounded continuous variable, while baseline tumor burden was modeled as the sum of target lesions in millimeters.

### Study dates, visits, and follow-up structure

A fixed study start date was used as the temporal anchor for simulation. Screening dates were generated by applying random offsets to the study start date, and randomization dates were assigned shortly thereafter. Treatment start date was aligned with randomization for treated patients. Withdrawal was simulated probabilistically, with withdrawal dates assigned only to patients meeting a simulated withdrawal criterion. Last-contact or last-alive dates were generated to support censoring in survival analyses.

Visit schedules were constructed to resemble a standard early oncology trial assessment calendar, including Screening, Baseline, Week 3, Week 6, Week 12, Week 18, Week 24, and then later visits at approximately 12-week intervals. Visit-date jitter was introduced to mimic realistic operational variability around nominal time windows. Each visit record included visit name, visit number, actual date, study day, cycle number where applicable, planned-visit flag, and window deviation.

### Treatment exposure simulation

Exposure was simulated at the dosing-event level. Treated patients were assigned investigational dosing records across repeated 21-day cycles, while control patients had no active investigational exposure by design. Dose levels were specified using fixed categories, and each record captured dose date, cycle, day within cycle, administered dose, dose unit, dose form, and indicators for administration, dose reduction, and dose interruption.

Dose modifications were linked probabilistically to adverse event burden, especially grade 3 or higher toxicity. As a result, exposure intensity varied across treated patients in a way intended to emulate real oncology dosing patterns, where treatment delivery is shaped by both protocol design and emerging toxicity. Patient-level pharmacokinetic summaries, including AUC_0_24, AUC_0_72, and Cmax, were then simulated as dose-dependent variables with additional log-normal inter-patient variability to support exploratory exposure-response analyses.

### Survival outcome generation

Patient-level survival outcomes were simulated using latent-risk-based exponential time-to-event models. For each patient, a latent risk score was calculated from standardized baseline variables including age, ECOG, baseline tumor burden, LDH, CRP, albumin, and ctDNA fraction. This approach was chosen because oncology survival risk is not driven by a single feature but by a weighted combination of disease burden, systemic inflammation, host functional status, and molecular disease activity (Petrelli et al., 2015; McMillan, 2013).

For overall survival, patient-specific hazard was defined as the product of a baseline hazard and an exponential function of the latent risk score and treatment effect. The treatment effect was specified as beneficial in the treatment arm, with additional benefit in biomarker-positive patients and partial attenuation in patients with ECOG 2. Event times were sampled from exponential distributions and then compared with independent censoring times. Observed overall survival duration was defined as the minimum of event and censoring time, and the overall survival event indicator was set to 1 when death occurred before censoring.

Progression-free survival was simulated similarly, but with a shorter baseline timescale than overall survival. Progression-free survival hazard included its own treatment coefficient and biomarker interaction. Event times reflected progression or death, whichever occurred first. Censoring occurred at last evaluable assessment or last known alive date when no progression or death had been observed. These simulated rules allowed progression-free survival to occur earlier and more frequently than overall survival, consistent with clinical oncology practice.

### Tumor response and longitudinal tumor burden modeling

Tumor response was simulated at both the categorical and continuous levels. First, each patient was assigned a probability of response using a logistic model incorporating treatment assignment, biomarker status, ECOG, and standardized baseline biomarker and disease-burden variables. This yielded a patient-specific responder probability, from which a binary responder status was sampled.

Best overall response categories were then assigned probabilistically according to responder status. Responders had greater probabilities of complete response or partial response, whereas non-responders were more likely to have stable disease or progressive disease. Objective response rate was defined as the proportion with complete or partial response, and clinical benefit rate as the proportion with complete response, partial response, or stable disease.

Longitudinal tumor burden was simulated as the sum of target lesions over time. For each patient, tumor burden at each assessment visit was modeled as a function of baseline tumor burden, a patient-specific slope, and random error. Responders in the treatment arm were assigned negative slopes to produce shrinkage over time, whereas control patients or non-responders were assigned flatter or positive slopes. Additional new-lesion and non-target progression flags were simulated to support progression logic and RECIST-like downstream analyses (Eisenhauer et al., 2009). Percent change from baseline was derived for waterfall and spider plot visualizations.

### Laboratory data generation

Longitudinal laboratory variables were simulated for each patient at each visit. The laboratory panel included ALT, AST, creatinine, hemoglobin, neutrophils, platelets, LDH, CRP, and albumin. Each analyte had a baseline distribution and a follow-up trend model intended to reflect either disease evolution, treatment toxicity, or both.

ALT and AST were generated with mild baseline variability and were allowed to rise in association with simulated hepatotoxicity. Creatinine was modeled with a narrow baseline distribution and small toxicity-linked changes. Hemoglobin, neutrophils, and platelets were allowed to decline modestly over time, particularly in patients with progressive disease or treatment-associated marrow suppression. Longitudinal LDH and CRP were modeled to track tumor burden and inflammatory progression, while albumin was modeled to decline with ongoing progression and systemic deterioration. Laboratory normal ranges and abnormality flags were carried alongside numeric values to support SDTM-like laboratory standardization and ADaM analysis derivations.

### Biomarker data generation

Longitudinal biomarker simulation focused on one principal continuous disease-monitoring biomarker, ctDNA fraction, together with exploratory inflammatory, immune, angiogenic, and proliferative markers. The biomarker panel included ctDNA fraction, IL-6, TNFα, IFNγ, VEGF, Ki-67 score, and PD-L1 score.

Baseline ctDNA fraction was generated as a bounded continuous variable, and post-baseline ctDNA values were modeled differently according to treatment arm and response status. Responders in the treatment arm showed steeper declines over time, non-responders in the treatment arm showed smaller declines or stabilization, and control or progressing patients showed increases over time. IL-6 and TNFα were modeled to rise in the presence of progression or systemic inflammatory activity. IFNγ was allowed to increase in immune-responsive or biomarker-positive treated patients. VEGF was linked loosely to tumor burden and progression, while Ki-67 and PD-L1 were simulated as exploratory markers of proliferation and immune context. Missingness was introduced at higher rates for biomarkers than for routine laboratories to reflect the operational reality of translational sampling in oncology trials.

### Adverse event generation

Adverse events were simulated at the event level rather than the patient-summary level. Each adverse event record included preferred term, system organ class, start date, end date, grade, serious flag, relatedness flag, action taken, outcome, adverse event of special interest flag, and death-related adverse event flag.

The probability of any adverse event and grade 3 or higher adverse event was modeled as a function of treatment assignment, ECOG, age, and baseline LDH. Event timing and duration were sampled from bounded distributions after treatment start. Preferred terms were sampled from a fixed oncology-style vocabulary including nausea, fatigue, anemia, neutropenia, diarrhea, rash, ALT increased, AST increased, infection, and decreased appetite. The treatment arm was enriched for several on-target or treatment-associated toxicities, including diarrhea, rash, transaminase increase, and neutropenia. Seriousness, relatedness, and treatment action were probabilistically linked to event grade and treatment arm. These event-level data were then used to derive patient-level safety flags such as any adverse event, grade 3 or higher adverse event, serious adverse event, and discontinuation due to toxicity.

### Concomitant medication generation

Concomitant medication episodes were simulated using a Poisson count model per patient. Medication classes included analgesics, antiemetics, antibiotics, corticosteroids, and proton pump inhibitors. Each record included medication term, class, start date, end date, and indication. Although these data were not central to the primary efficacy findings, they were included to preserve the clinical-trial realism of the raw and SDTM-like datasets.

### Missing-data mechanism

Missingness was introduced to reflect practical trial conduct. Baseline variables had low rates of missingness, whereas longitudinal laboratory, tumor, and biomarker assessments had progressively higher missingness rates. Missingness was increased after simulated progression, withdrawal, or death so that later time points were less likely to be observed in clinically deteriorating patients. This approach was used to emulate the informative and operationally driven incompleteness common in oncology trial data.

### Dataset construction

The workflow generated raw source datasets, SDTM-like standardized domains, and ADaM-like analysis-ready datasets. Raw datasets included demographics_raw.csv, visits_raw.csv, exposure_raw.csv, biomarker_raw.csv, labs_raw.csv, tumor_raw.csv, adverse_events_raw.csv, conmeds_raw.csv, and survival_raw.csv. SDTM-like datasets included DM, SV, EX, LB, TU, AE, and CM domains. These standardized records used clinical-trial-style naming conventions for identifiers, timing variables, baseline descriptors, and event descriptors. ADaM-like datasets included ADSL, ADTTE, ADLB, ADTU, ADBM, and ADAE. ADSL contained one record per patient and included treatment flags, baseline variables, exposure summaries, response variables, time-to-event variables, and patient-level safety flags. ADTTE contained one record per patient per time-to-event parameter, including OS, PFS, TTD, and TTAE. ADLB, ADTU, and ADBM stored longitudinal laboratory, tumor, and biomarker records with derived baseline, change, and percent change variables. ADAE contained analysis-ready event-level safety records, including first-occurrence and treatment-emergent flags.

### Derived variables

Core derived variables were generated according to standard oncology analysis logic. In ADSL, overall survival days were defined as the difference between death or censor date and randomization date plus one day. Progression-free survival days were defined analogously using progression, death, or censor date. Censoring indicators distinguished event from censored observations. Response flags classified patients with complete or partial response as responders, while clinical benefit included complete response, partial response, and stable disease. Dose intensity was defined as actual cumulative dose divided by planned cumulative dose. Safety flags identified any grade 3 or higher adverse event, any serious adverse event, and discontinuation due to toxicity. A biomarker-high flag was derived from the baseline continuous biomarker distribution to support subgroup-style analyses.

In ADLB, ADTU, and ADBM, baseline, change from baseline, and percent change from baseline were calculated for each patient-parameter combination. These derivations supported both summary tables and longitudinal visualization.

### Statistical analysis

All analyses were descriptive or exploratory because the dataset was synthetic. Baseline characteristics were summarized by treatment arm using counts, percentages, and means. Best overall response, objective response rate, and clinical benefit rate were calculated from subject-level response categories. Exposure metrics were summarized at both patient and arm levels.

Time-to-event analyses used Kaplan-Meier estimators for overall survival and progression-free survival, with treatment-arm comparisons assessed using approximate hazard ratios from Cox proportional hazards models and p-values from log-rank testing where estimable. Subgroup hazard-ratio exploration was performed across selected baseline strata. Longitudinal tumor and biomarker behavior were summarized through change-from-baseline statistics and visualized using mean trajectory plots, waterfall plots, and spider plots. Safety was summarized at the patient level and adverse event term level. Exposure-response analysis examined the association between simulated pharmacokinetic metrics and progression-free survival among treated patients.

### Tables, figures, and decision framework

The analytical pipeline produced publication-style outputs including baseline characteristic tables, treatment exposure summaries, best overall response tables, adverse event summaries, laboratory and biomarker tables, Kaplan-Meier plots, subgroup forest plots, tumor-response plots, biomarker trajectory plots, and exposure-response plots. The final layer of the workflow translated these outputs into a simple decision-oriented interpretation of whether the simulated treatment demonstrated a credible efficacy signal with manageable toxicity and biomarker support. This final decision layer was included because in real oncology development, the purpose of integrated clinical analysis is not only to describe data, but to support rational go/no-go or enrichment decisions.

### Software and reproducibility

The full workflow was implemented in Python environment. Data simulation, transformation, and analysis used standard scientific computing libraries including NumPy, pandas, Matplotlib, and lifelines. The notebook generated all datasets, analyses, tables, figures, and decision summaries in a fully reproducible sequence. Because the dataset was synthetic and no real human participants were involved, ethical approval and informed consent were not required. All datasets and models used in this study are openly available in the GitHub repository using this link: https://github.com/mpetalcorin/synthetic-oncology-clinical-trial-framework.

## Results

### Treatment arms entered the study with broadly comparable baseline characteristics

The synthetic phase II cohort included 220 randomized patients, 103 assigned to control and 117 assigned to treatment (Supplementary Table S1). Baseline demographic and clinical features were well balanced between arms. Mean age was 62.7 years in the control arm and 61.5 years in the treatment arm. Female patients comprised 43 of 103 control patients and 52 of 117 treated patients. ECOG performance status distributions were also similar, with ECOG 0, 1, and 2 observed in 32, 52, and 19 control patients, respectively, compared with 39, 51, and 27 treated patients. Baseline tumor burden was comparable between arms, with mean values of 101.4 mm in the control arm and 98.6 mm in the treatment arm. Likewise, baseline LDH, CRP, albumin, and ctDNA fraction showed no major imbalances, and biomarker-positive disease was present in 44 control patients and 48 treated patients (Supplementary Table S1). These findings indicate that the simulated randomization process generated treatment groups suitable for comparative efficacy and safety assessment.

### Treatment generated meaningful exposure with realistic dose modification patterns

Treatment exposure analyses showed that the investigational arm achieved measurable drug delivery while preserving clinically plausible dose modification patterns (Supplementary Table S2). Among treated patients, the mean cumulative dose was 527.6 mg and the mean dose intensity was 0.779. Dose modifications occurred frequently enough to resemble real oncology development datasets, with 85 dose reductions and 69 dose interruptions recorded in the treatment arm. As expected by design, the control arm had no active drug exposure, with a cumulative dose mean of 0.0 mg. These results demonstrate that the dataset captured not only treatment assignment, but also operationally relevant variability in how therapy was actually delivered.

### Treatment improved best overall response and increased clinical benefit

Antitumor activity favored the treatment arm (Supplementary Table S3). In the control arm, best overall response consisted of 3 complete responses, 24 partial responses, 37 stable disease events, and 39 progressive disease events, yielding an objective response rate of 26.2% and a clinical benefit rate of 62.0%. In the treatment arm, best overall response included 3 complete responses, 40 partial responses, 42 stable disease events, and 32 progressive disease events, corresponding to an objective response rate of 36.8% and a clinical benefit rate of 72.4%. These results show that treatment increased the proportion of patients achieving tumor shrinkage and increased the proportion deriving disease control.

### Treatment reduced tumor burden over time and shifted patient-level tumor trajectories

Longitudinal tumor analyses confirmed the response advantage. Mean percent change from baseline in tumor burden favored the treatment arm across follow-up visits (Figure 2), indicating greater group-level tumor shrinkage and less progressive growth than observed in the control arm. The waterfall plot further showed that the treatment arm contained many of the deepest tumor reductions, with top responders exhibiting marked declines in tumor burden, including percent changes approaching -98% (Figure 3). The spider plot reinforced this pattern by showing that treated patients more often followed sustained downward or stabilized trajectories, whereas control patients more frequently demonstrated rising tumor burden over time (Figure 4). Together, these tumor analyses show that treatment improved both average tumor dynamics and the distribution of individual radiographic responses.

**Figure 1.**
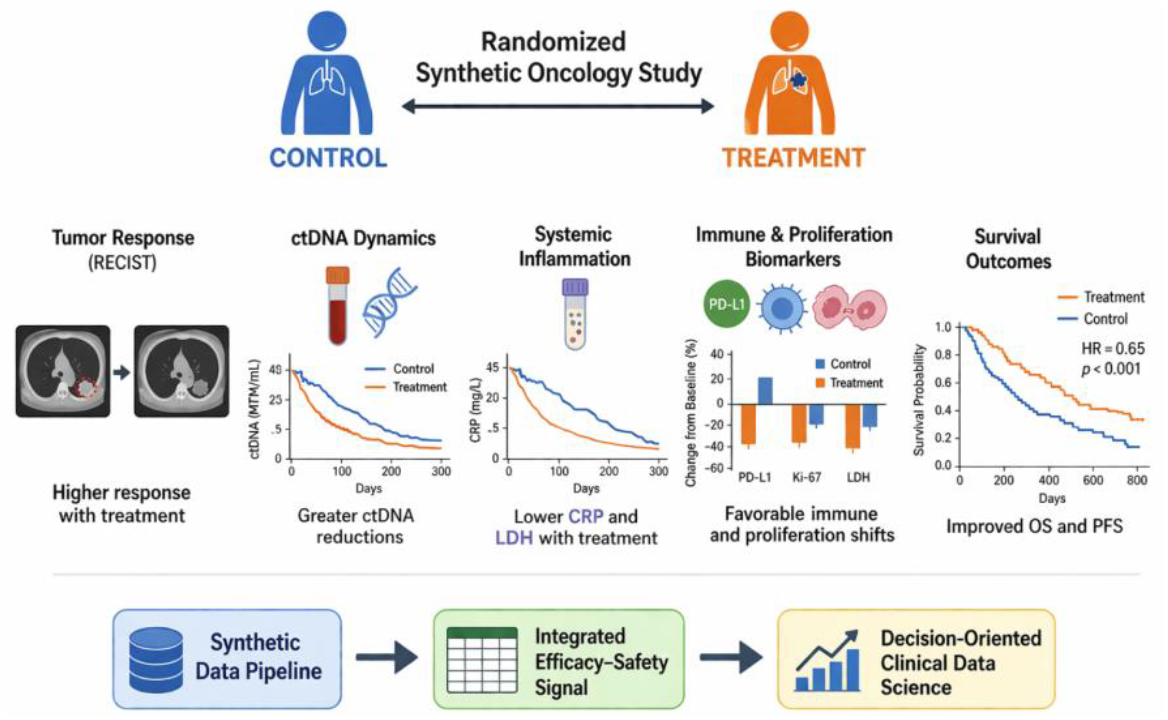
Conceptual overview of the synthetic oncology clinical trial framework and integrated efficacy-safety findings. This schematic illustrates the overall design logic and main findings of the synthetic, literature-informed oncology study. Patients were randomized to control or treatment arms at the top of the workflow. The middle layers summarize the major analytic dimensions captured in the manuscript, including radiographic tumor response, circulating tumor DNA dynamics, systemic inflammation, immune and proliferation biomarkers, and survival outcomes. The lower portion of the figure highlights the central interpretation of the study, namely that a synthetic data pipeline can generate an integrated efficacy-safety signal across layers and support decision-oriented clinical data science.

**Figure 2.**
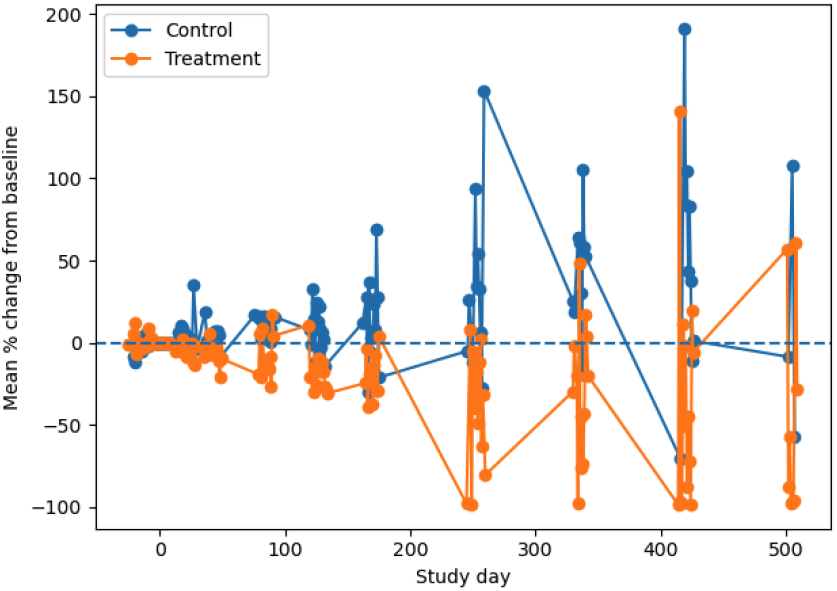
Mean tumor percent change from baseline over time by treatment arm. Mean percent change in tumor burden from baseline is shown across study visits for the control and treatment arms. Negative values indicate tumor shrinkage, whereas positive values indicate tumor growth. The figure summarizes group-level radiographic dynamics during follow-up.

**Figure 3.**
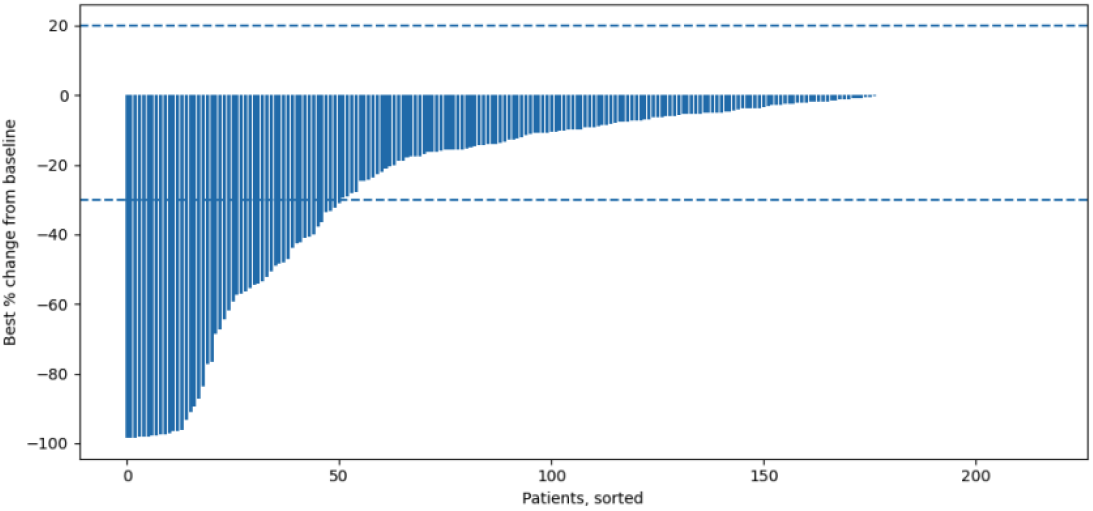
Waterfall plot of best tumor response. Waterfall plot of best percent change from baseline in tumor burden for individual patients, ordered from greatest decrease to greatest increase. Many patients crossed the conventional partial-response threshold, and only a minority showed clear growth.

**Figure 4.**
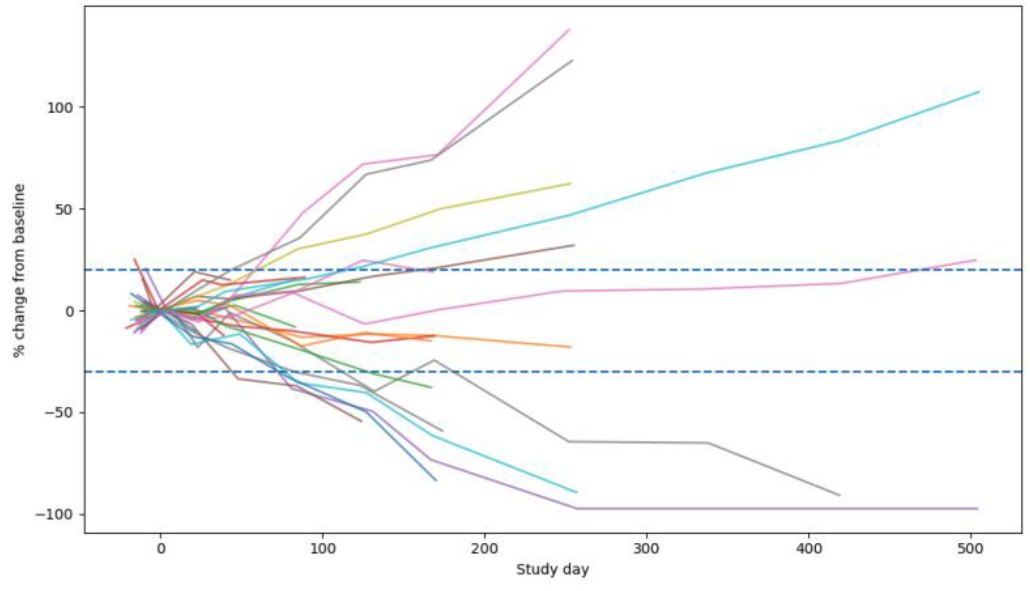
Spider plot of longitudinal tumor trajectories. Spider plot showing longitudinal percent change from baseline in tumor burden for representative patients over time. Each line represents one patient trajectory. The plot illustrates both sustained shrinkage in some patients and progression in others.

### Treatment prolonged overall survival and progression-free survival

Time-to-event analyses showed a clear efficacy signal in favor of treatment. Median overall survival increased from 135 days in the control arm to 288 days in the treatment arm (Figure 5, Supplementary Table S4). The approximate hazard ratio for overall survival was 0.661 (95% CI, 0.480-0.911; p = 0.011), indicating a lower risk of death in the treatment arm. Progression-free survival also improved, with median progression-free survival increasing from 116 days in the control arm to 208 days in the treatment arm (Figure 6, Supplementary Table S4). The approximate hazard ratio for progression-free survival was 0.601 (95% CI, 0.418-0.864; p = 0.006). These results show that the simulated treatment effect translated into both delayed progression and prolonged survival.

**Figure 5.**
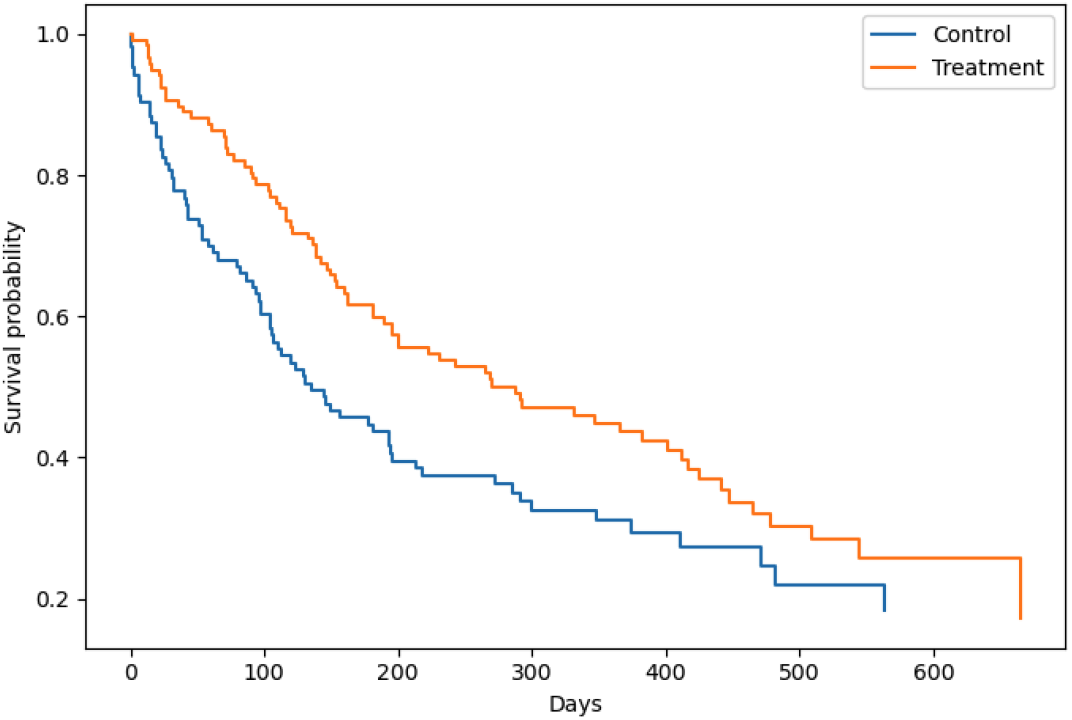
Kaplan-Meier overall survival by treatment arm. Kaplan-Meier estimates of overall survival are shown for the control and treatment arms in the synthetic phase II oncology cohort. Overall survival was defined as the time from randomization to death from any cause. Patients without an observed death event were censored at the last known alive date. The separation between the curves illustrates the simulated survival benefit in the treatment arm relative to control.

**Figure 6.**
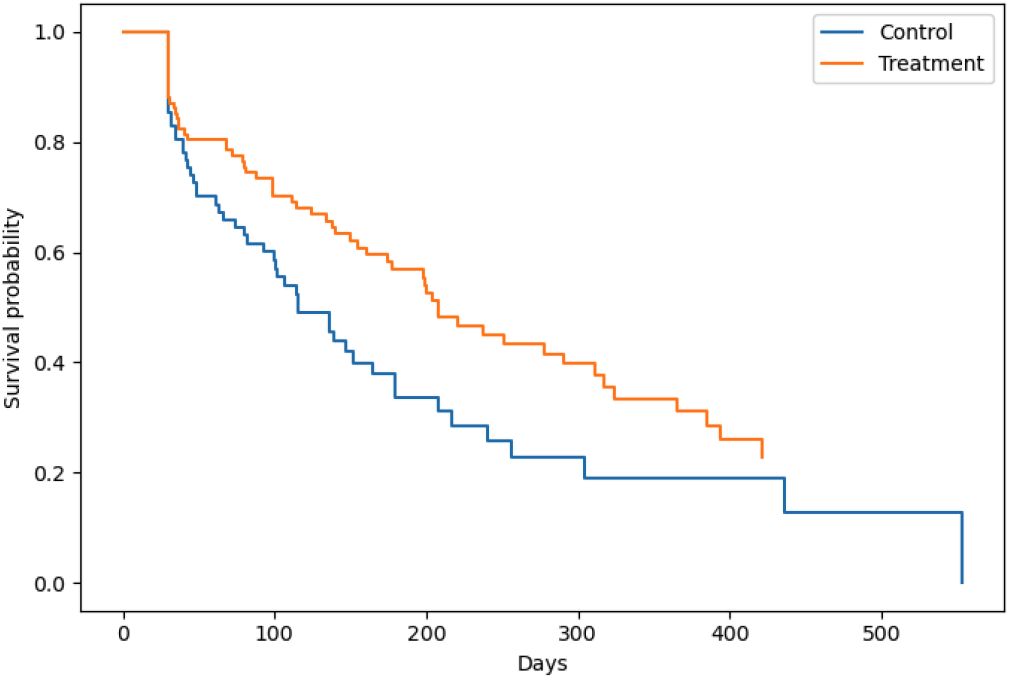
Kaplan-Meier progression-free survival by treatment arm. Kaplan-Meier estimates of progression-free survival are shown for the control and treatment arms. Progression-free survival was defined as the time from randomization to the first documented disease progression or death, whichever occurred first. The treatment arm demonstrates delayed progression relative to control in the simulated dataset.

### Treatment maintained directional benefit across major subgroups

Exploratory subgroup analyses suggested that the survival benefit remained directionally consistent across several clinically relevant strata (Figure 7, Supplementary Table S5). Among male patients, the overall survival hazard ratio was 0.631 (95% CI, 0.414-0.960; p = 0.032), whereas among female patients the hazard ratio was 0.709 (95% CI, 0.432-1.164; p = 0.174). In older patients, defined as those at or above the median age, the hazard ratio was 0.648 (95% CI, 0.433-0.968; p = 0.034), while in younger patients it was 0.700 (95% CI, 0.412-1.189; p = 0.187). In the ECOG 0 subgroup, the hazard ratio was 0.852 (95% CI, 0.479-1.514; p = 0.584). Although several subgroup estimates were imprecise, the overall pattern remained broadly treatment-favorable, supporting consistency of effect rather than a signal confined to a single stratum.

**Figure 7.**
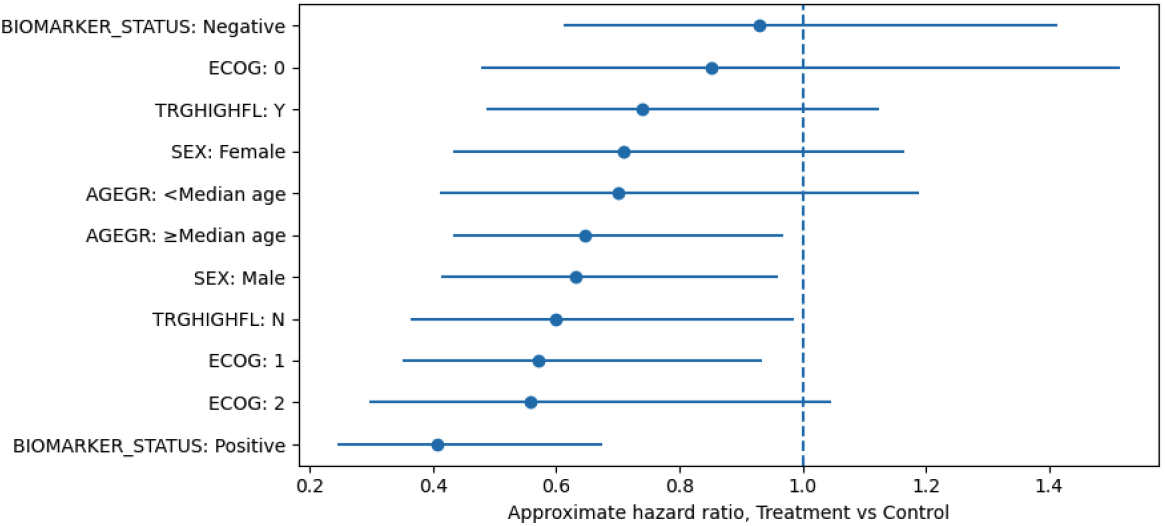
Forest plot of overall survival subgroup hazard ratios. Forest plot of subgroup-specific hazard ratios for overall survival comparing treatment versus control. Points indicate hazard ratio estimates and horizontal lines indicate 95% confidence intervals. The vertical reference line at 1.0 indicates no difference between treatment arms.

### Treatment shifted ctDNA and other biomarkers in biologically coherent directions

Biomarker analyses showed treatment-associated changes that aligned with the efficacy findings (Supplementary Table S6, Figure 8). In the control arm, mean ctDNA fraction increased by 0.011 from baseline, corresponding to a mean percent increase of 94.2%. In contrast, the treatment arm demonstrated a more favorable ctDNA profile over time in the visual longitudinal analysis (Figure 8), consistent with stronger disease control. Additional biomarker trends also supported biologically coherent treatment effects. Mean IFNγ increased by 1.775 in the treatment arm compared with 0.053 in the control arm, while Ki-67 score decreased by 0.599 in the treatment arm but increased slightly by 0.196 in the control arm. IL-6 and TNFα rose in both arms, but the ctDNA and proliferative marker patterns were more consistent with treatment benefit. These findings support the inclusion of both binary biomarker status and continuous ctDNA measurements in the analytic framework.

**Figure 8.**
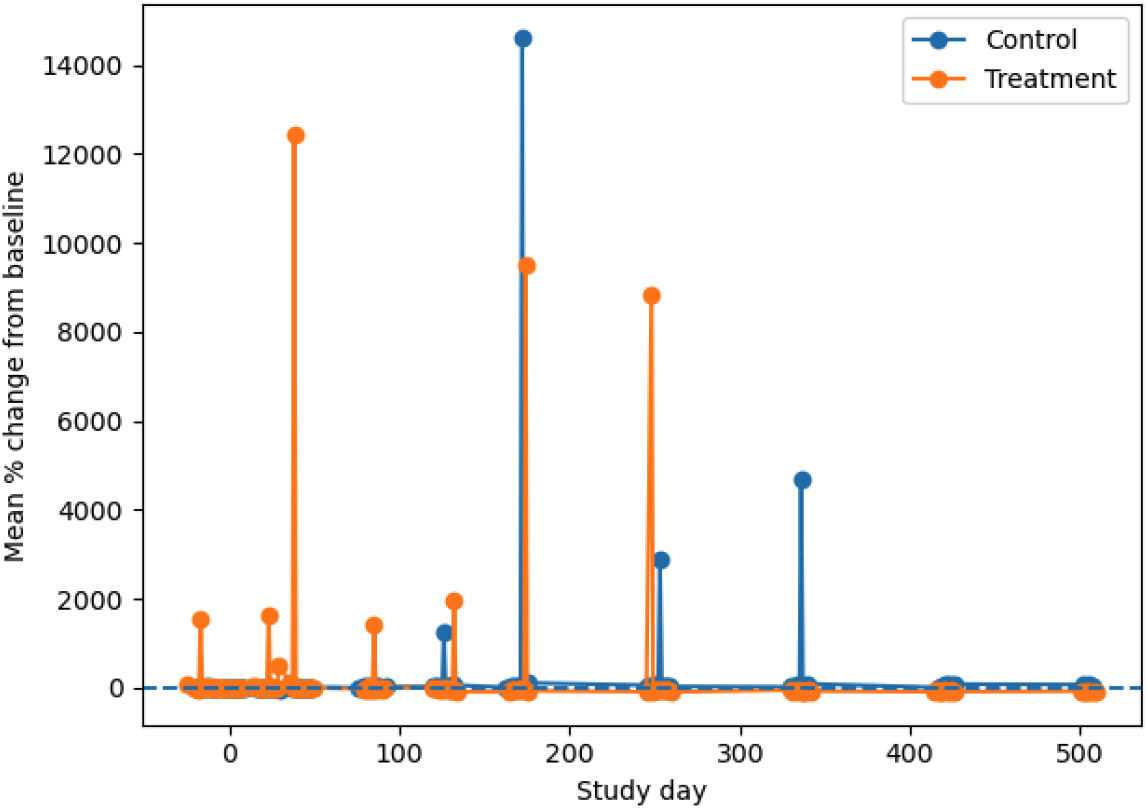
Change in circulating tumor DNA over time by treatment arm. Mean percent change from baseline in circulating tumor DNA is shown over time for the control and treatment arms. Despite variability and several extreme values, the overall treatment trajectory was more favorable than control.

### Exposure-response analysis revealed only a weak association between exposure and progression-free survival

Exploratory pharmacometric analysis showed that higher treatment exposure did not strongly translate into longer progression-free survival in this synthetic cohort (Figure 9). The correlation between AUC_0_24 and progression-free

**Figure 9.**
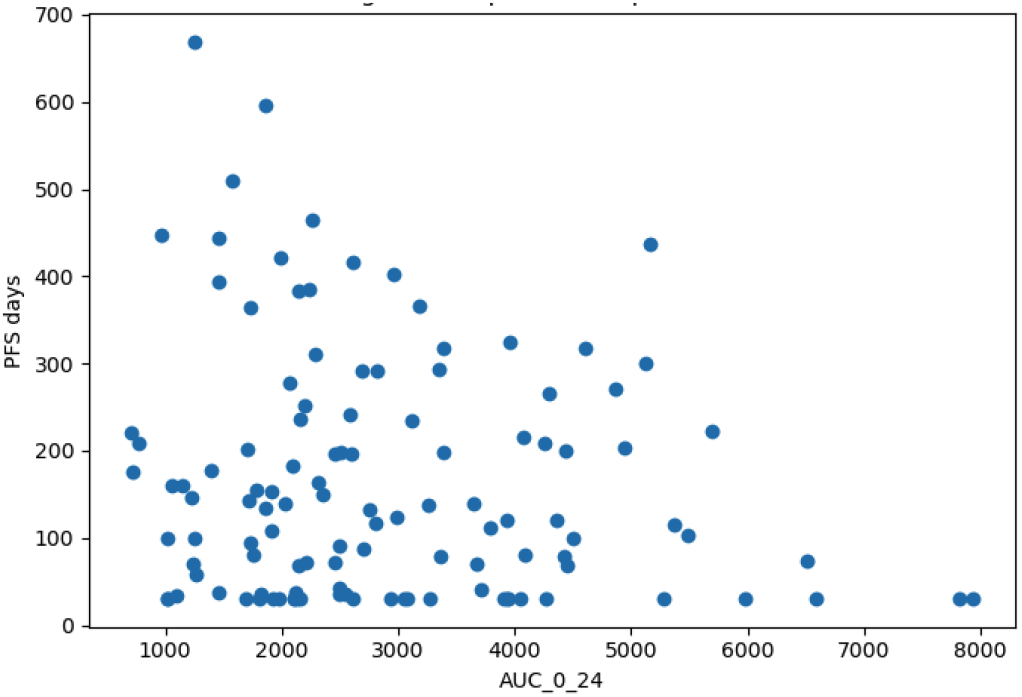
Exposure-response relationship between drug exposure and progression-free survival. Scatter plot showing the relationship between AUC_0_24 and progression-free survival among treated patients. The broad spread indicates that exposure alone did not fully account for efficacy heterogeneity in this synthetic cohort.

### Treatment altered the safety profile without overwhelming tolerability

Safety analyses showed that the treatment arm produced a distinct but manageable toxicity profile (Supplementary survival duration was modest and negative (r = −0.160). This weak relationship suggests that within the simulated exposure range, efficacy was not driven solely by drug exposure magnitude, and that disease biology and baseline risk remained important determinants of outcome. Even so, the analysis demonstrated that the project can connect dosing, pharmacokinetics, and efficacy in a format familiar to translational oncology development.

Table S7, Figure 10). Any adverse event occurred in 94.2% of control patients and 91.5% of treated patients. Grade 3 or higher adverse events occurred in 50.5% of control patients and 46.2% of treated patients. Serious adverse events were more frequent with treatment, occurring in 25.6% of treated patients compared with 17.5% of controls. Related adverse events were also more common in the treatment arm, 76.9% versus 59.2%, and discontinuation due to adverse events occurred in 6.8% of treated patients compared with 2.9% of controls (Supplementary Table S7). Event-term distributions showed that ALT increased, AST increased, rash, and diarrhea were more frequent in treated patients, whereas anemia and neutropenia were somewhat more frequent in control patients in this simulated run (Figure 10). Overall, treatment increased treatment-related toxicity, but did not produce an implausible safety burden that would negate the observed efficacy advantages.

**Figure 10.**
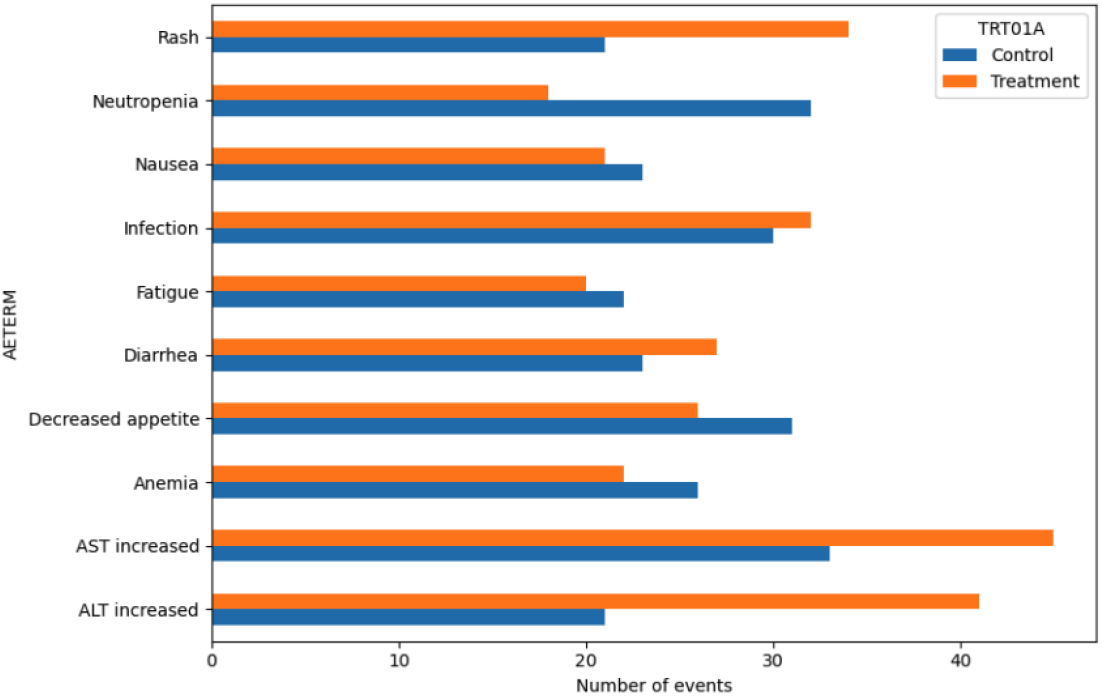
Adverse-event frequency by treatment arm. Horizontal bar chart showing the frequency of adverse-event records by preferred term and treatment arm. ALT increased, AST increased, rash, infection, and diarrhea were more frequent in the treatment arm, whereas other events were mor

### Laboratory trends tracked both disease activity and treatment effects

Laboratory summaries captured clinically interpretable biological changes over time (Supplementary Table S8). In the control arm, mean CRP increased by 0.373, whereas albumin decreased by 0.054 and hemoglobin decreased by 0.305. In the treatment arm, albumin also declined modestly, by 0.073, while ALT increased by 0.343 and AST changed by −0.231 on average. LDH remained relatively stable in both groups, with a mean change of 0.630 in control patients. These laboratory patterns complement the efficacy and safety findings by showing that the dataset supports simultaneous monitoring of inflammation, organ function, hematologic effects, and disease-related biology.

### The integrated pipeline produced decision-ready evidence supporting further study

Across the full workflow, the project successfully translated patient-level simulation into structured data, derived datasets, interpretable analyses, and publication-style outputs. Baseline comparability (Supplementary Table S1), exposure characterization (Supplementary Table S2), tumor response improvement (Supplementary Table S3, Figures 2-4), survival benefit (Figures 5-7, Supplementary Table S4), biomarker shifts (Supplementary Table S6, Figure 8), and manageable toxicity (Supplementary Table S7, Figure 10) together support a coherent development narrative. The executed notebook’s final integrated summary concluded that the treatment arm showed a credible efficacy signal with manageable toxicity, thereby supporting progression to a larger confirmatory or biomarker-enriched study. In that sense, the workflow achieved its intended sequence of Patient → Data → Dataset → Analysis → Tables/Figures → Decision, and generated a decision-ready oncology clinical data science demonstration.

## Discussion

The central finding of this synthetic oncology study is not simply that the treatment arm performed better than control across several endpoints, but that the pattern of benefit was internally coherent across multiple biological layers. In the executed computational simulations, treatment improved best overall response, shifted longitudinal tumor trajectories downward, reduced ctDNA more favorably, and prolonged both progression-free survival and overall survival. That constellation matters because, in real oncology, efficacy rarely rests on one endpoint alone. Rather, convincing antitumor activity emerges when imaging, blood-based biomarkers, and time-to-event outcomes move in the same direction. At the systems level, that coherence suggests the simulated data-generating process captured a plausible coupling between tumor-cell kill, reduced net tumor mass, altered inflammatory signaling, and delayed clinical deterioration. This is conceptually aligned with modern oncology frameworks in which tumor control depends on both direct effects on cancer-cell fitness and indirect effects on host immunity, stromal interaction, and systemic inflammatory state (Chen & Mellman, 2013; Doroshow et al., 2021).

Among the biomarker findings, the ctDNA signal is especially important because it provides a mechanistic bridge between molecular events and clinical outcomes. ctDNA is a blood-borne representation of tumor-derived nucleic acids released during tumor-cell turnover, including apoptosis and necrosis, and it can capture temporal and spatial heterogeneity more dynamically than a single tissue biopsy. For that reason, early falls in ctDNA can precede or complement radiographic response, whereas persistent or rising ctDNA can indicate inadequate cytoreduction, clonal persistence, or early progression. Multiple recent studies have shown that early ctDNA dynamics can correlate with radiographic response and long-term outcomes, and that longitudinal ctDNA modeling can improve survival prediction in advanced solid tumors, including randomized trial settings (Liu et al., 2020; Kansara et al., 2023; Assaf et al., 2023).

That is why the ctDNA findings in this computational modeling are biologically meaningful. In the treatment arm, ctDNA moved in a more favorable direction than in control, and that direction matched the improved tumor-response and survival curves. Scientifically, that pattern implies a lower effective residual malignant burden under treatment. At the molecular level, reduced ctDNA is compatible with fewer actively proliferating clones, less tumor mass available for shedding DNA fragments into plasma, and slower expansion of resistant subclones. At the systems level, serial ctDNA acts as a compressed summary of tumor ecology over time, integrating treatment sensitivity, tumor turnover, and evolutionary adaptation. In a flagship aAidea-style project, this is valuable because it demonstrates how a continuous biomarker can enrich classical RECIST-like endpoints rather than merely duplicate them.

The LDH, CRP, and albumin findings add an important second layer of interpretation. Tumor burden alone does not determine patient outcome. Cancer progression is also shaped by metabolic stress, tissue injury, cachexia, host inflammatory response, and organ reserve. LDH is especially relevant because it sits close to the metabolic core of aggressive cancer behavior. Elevated circulating LDH often reflects high glycolytic flux, tissue turnover, hypoxia-associated stress, and large tumor burden, and high LDH has been repeatedly associated with worse survival across solid tumors (Petrelli et al., 2015). CRP and albumin capture another part of the same system. CRP rises with systemic inflammation, while low albumin reflects a combination of inflammation, impaired nutritional state, altered hepatic protein synthesis, and advanced disease biology. Inflammation-based composite frameworks such as the Glasgow Prognostic Score, and CRP-to-albumin ratio approaches, have shown reproducible prognostic value across cancers (McMillan, 2013; Shibutani et al., 2016).

This helps explain why the simulated dataset becomes more convincing when these markers move coherently with efficacy. A treatment can shrink tumors but still fail to deliver meaningful survival if systemic inflammation remains high, if metabolic stress persists, or if the host enters progressive functional decline. Conversely, when treatment-associated tumor control is accompanied by favorable biomarker movement in ctDNA and stabilization of inflammation-related variables, the survival signal becomes more believable. At a systems level, these markers function as an interface between tumor-intrinsic biology and organism-level physiology. They remind us that cancer is not only a localized mass problem, but also a whole-body inflammatory and metabolic disease state.

The exploratory biomarker panel, including IFNγ, PD-L1, VEGF, and Ki-67, is useful because it points toward distinct biological axes of response. IFNγ is central to antitumor immunity, particularly to T-cell activation, antigen presentation, and macrophage polarization, although its effects can be context dependent and can also shape immune resistance pathways. PD-L1, meanwhile, remains an imperfect but still widely used biomarker of response to immune-checkpoint blockade. Its value lies not in universal predictiveness, but in its role as one component of a broader immune context that includes inflammatory signaling, immune-cell infiltration, and tumor mutational architecture. VEGF reflects angiogenic drive and abnormal vascular signaling, both of which influence nutrient delivery, hypoxia, immune exclusion, and therapeutic penetration. Ki-67 reflects proliferative state and thus the tempo of tumor-cell cycling (Jorgovanovic et al., 2020; Doroshow et al., 2021; Tugues et al., 2011; Menon et al., 2019).

In the context of executed simulations, these biomarkers are read less as standalone determinants and more as a networked panel. A fall in Ki-67-like proliferative activity together with favorable ctDNA behavior suggests reduced net clone expansion. A rise in IFNγ may suggest a more inflamed, treatment-responsive immune environment, although interpretation depends on context. VEGF-related signals can help explain whether nonresponse is tied to angiogenic escape, abnormal vasculature, or microenvironmental support for persistence. PD-L1 can help stratify immune context, but may not be overinterpreted in isolation because both biological heterogeneity and assay variability limit its standalone reliability. Scientifically, this is the value of a systems-level analysis, the markers are strongest when interpreted together, not separately.

IL-6 deserves special attention because it is one of the clearest molecular links between cancer-cell behavior, host inflammation, and therapeutic resistance. IL-6 signaling has been implicated in proliferation, survival, angiogenesis, invasion, metastasis, metabolic rewiring, and resistance to therapy. In practical terms, persistent IL-6 activity can sustain a tumor-supportive ecosystem even when radiographic shrinkage is observed early. That means a patient can show short-term tumor reduction yet still relapse quickly if inflammatory and resistance-promoting circuits remain active. This is one reason why dynamic biomarker panels are more informative than one-time measurements. They can distinguish transient tumor shrinkage from deeper biological reprogramming (Kumari et al., 2016).

Within a synthetic framework, IL-6 complements CRP and albumin as part of a multiscale inflammatory axis. At the molecular level, IL-6 can feed STAT3-centered transcriptional programs that enhance survival and adaptation. At the tissue level, it contributes to a permissive microenvironment. At the organism level, it supports systemic inflammatory burden. Therefore, if future versions of the project strengthen the longitudinal coupling among IL-6, CRP, albumin, and ctDNA, the simulation could more explicitly reproduce the biology of patients who initially respond but later develop inflammatory escape and progression.

The weak exposure-response relationship in the executed simulations should not automatically be interpreted as a flaw. In real clinical pharmacology, exposure is only one determinant of outcome. Once drug concentrations enter a pharmacologically active range, outcome can become more dependent on tumor biology, target engagement, bypass signaling, immune context, and baseline disease aggressiveness than on further exposure increases alone. This is especially true in oncology when resistant subclones, angiogenic escape, microenvironmental buffering, or systemic inflammation dominate the trajectory of disease. Thus, the modest exposure-response pattern in computational simulations is scientifically plausible, particularly in a heterogeneous cohort. It suggests that the synthetic trial is not overfitted to a simplistic assumption that “more drug always means more benefit,” and that is a strength in a translational dataset meant to resemble clinical reality. The biology of angiogenic signaling and immune context further supports this principle, because resistance and persistence can emerge despite adequate drug delivery (Tugues et al., 2011; Doroshow et al., 2021).

The real value of this project is methodological. It converts patient-level features into raw data, standardized datasets, analysis-ready datasets, integrated analyses, and decision-oriented outputs. That mirrors how oncology development actually works. The pipeline does not ask whether one biomarker is interesting in isolation. It asks whether baseline factors, longitudinal biomarkers, survival, exposure, safety, and subgroup structure together support a credible development decision. That is exactly the difference between a descriptive simulation and a drug-development modeling.

The main limitation is equally important to be kept in mind. This remains a synthetic, literature-informed dataset, not a real trial. Therefore, the discussion can address biological plausibility, internal coherence, and methodological utility, but it cannot claim clinical efficacy or true biomarker validation. The proper interpretation is that the computational simulation reproduces a believable oncology signal architecture, one that is consistent with published biology and clinical-trial behavior, and therefore serves as a strong testbed for clinical data science, translational modeling, and clinical trial study presentation. Future versions would become even stronger by calibrating the simulation against disease-specific trial distributions, explicitly modeling resistance emergence, and incorporating joint models that couple ctDNA, imaging, and survival more tightly, as has already been demonstrated in longitudinal ctDNA survival work (Kansara et al., 2023; Assaf et al., 2023).

## Conclusion

In this work, we developed a synthetic, literature-informed oncology clinical trial framework that reproduces the logic of modern cancer drug development from patient-level simulation to decision-oriented interpretation. The full workflow connected baseline patient features, longitudinal tumor measurements, ctDNA dynamics, laboratory and exploratory biomarkers, exposure records, safety events, and survival outcomes through structured raw datasets, SDTM-like domains, and ADaM-like analysis datasets. This design allowed the study to move beyond isolated endpoint simulation and instead generate a coherent efficacy-safety narrative across multiple biological and clinical layers.

The executed analyses showed that the treatment arm achieved greater tumor response, more favorable tumor trajectories, longer progression-free survival, longer overall survival, and more favorable ctDNA dynamics than the control arm. Importantly, these benefits emerged together rather than in isolation. That internal coherence makes the synthetic dataset more scientifically credible because it reflects the multiscale nature of cancer therapeutics, where molecular response, radiographic change, and survival outcome should generally align if a treatment is truly biologically active. At the same time, the treatment arm also showed increased treatment-related toxicity, preserving the realistic trade-off between efficacy and safety that characterizes real oncology development programs.

At the molecular level, the framework highlights the importance of integrating tumor burden, ctDNA, inflammatory markers, and exploratory immune and proliferative signals when interpreting treatment effect. At the systems level, it shows that decision-making in oncology depends on linking patient heterogeneity, exposure, efficacy, safety, and biomarker evolution into a unified analytical structure. This is the practical value of the Patient → Data → Dataset → Analysis → Tables/Figures → Decision model. It transforms a complex set of simulated clinical observations into decision-ready evidence that resembles the output expected in translational and pharmaceutical oncology settings.

The principal limitation is that all findings are based on synthetic rather than real patient-level data. Accordingly, the study should not be interpreted as evidence for any actual therapeutic agent or biomarker. Its value lies instead in methodological rigor, biological plausibility, reproducibility, and presentation. As such, this framework provides a strong prototype for clinical trial study development, internal analytics demonstration, dashboard design, biomarker strategy modeling, and training in clinical trial data science. Future extensions could calibrate the simulation to disease-specific trial distributions, include explicit resistance-emergence models, and implement more advanced joint modeling of imaging, ctDNA, and survival. Even in its present form, however, the study demonstrates that a well-designed synthetic oncology trial can serve as a powerful platform for developing and communicating end-to-end clinical data science in cancer research.

## Data Availability

All data produced are openly available online at https://github.com/mpetalcorin/synthetic-oncology-clinical-trial-framework.

## Supplementary Tables

**Supplementary Table S1.** Baseline demographic and disease characteristics by treatment arm.

| TRT01A | N | AGE MEAN | FEMALE N | ECOG 0 N | ECOG 1 N | ECOG 2 N | TUMORBL MEAN | LDHBL MEAN | CRPBL MEAN | ALBBL MEAN | CTDNABL MEAN | BIOMPOSS N |
| --- | --- | --- | --- | --- | --- | --- | --- | --- | --- | --- | --- | --- |
| Control | 103 | 62.747 | 43 | 32 | 52 | 19 | 101.361 | 254.022 | 12.985 | 3.882 | 0.141 | 44 |
| Treatment | 117 | 61.453 | 52 | 39 | 51 | 27 | 98.644 | 267.065 | 12.141 | 3.947 | 0.143 | 48 |

Baseline demographic and clinical characteristics are summarized by randomized treatment arm. Continuous variables are presented as summary statistics, and categorical variables as counts and percentages. The table demonstrates baseline comparability between groups.

**Supplementary Table S2.** Treatment exposure and dose intensity by treatment arm.

| TRT01A | N | cumulative dose mean | dose intensity mean | dose reductions total | dose interruptions total |
| --- | --- | --- | --- | --- | --- |
| Control | 92 | 0.0 |  | 0 | 0 |
| Treatment | 114 | 527.631 | 0.779 | 85 | 69 |

Treatment exposure summaries are presented by arm, including cumulative dose, dose intensity, dose reductions, and dose interruptions. These metrics provide context for interpretation of both efficacy and safety outcomes.

**Supplementary Table S3.** Best overall response by treatment arm.

| TRT01A | CR | PD | PR | SD | ORR | CBR |
| --- | --- | --- | --- | --- | --- | --- |
| Control | 3 | 39 | 24 | 37 | 0.262 | 0.620 |
| Treatment | 3 | 32 | 40 | 42 | 0.367 | 0.724 |

Best overall response categories are summarized by treatment arm, including complete response, partial response, stable disease, and progressive disease. Objective response rate and clinical benefit rate are also shown.

**Supplementary Table S4.** Survival summary for overall survival and progression-free survival.

| endpoint | control median | treatment median | hr | lcl | ucl | p value |
| --- | --- | --- | --- | --- | --- | --- |
| OS | 135.0 | 288.0 | 0.661 | 0.480 | 0.911 | 0.011 |
| PFS | 116.0 | 208.0 | 0.601 | 0.418 | 0.864 | 0.006 |

Overall survival and progression-free survival are summarized by treatment arm, together with approximate hazard ratios, confidence intervals, and p-values where estimable.

**Supplementary Table S5.** Subgroup hazard ratios for overall survival.

| subgroup | level | n | events | hr | lcl | ucl | p value |
| --- | --- | --- | --- | --- | --- | --- | --- |
| SEX | Male | 125 | 87 | 0.631 | 0.414 | 0.960 | 0.032 |
| SEX | Female | 95 | 63 | 0.709 | 0.432 | 1.1642 | 0.174 |
| AGEGR | ≥Median age | 123 | 95 | 0.647 | 0.433 | 0.968 | 0.034 |
| AGEGR | <Median age | 97 | 55 | 0.700 | 0.412 | 1.188 | 0.187 |
| ECOG | 0 | 71 | 47 | 0.852 | 0.479 | 1.514 | 0.584 |
| ECOG | 1 | 103 | 64 | 0.577 | 0.350 | 0.934 | 0.026 |
| ECOG | 2 | 46 | 39 | 0.558 | 0.298 | 1.046 | 0.069 |
| BIOMARKER_STATUS | Negative | 128 | 89 | 0.930 | 0.613 | 1.412 | 0.734 |
| BIOMARKER_STATUS | Positive | 92 | 61 | 0.407 | 0.245 | 0.674 | 0.001 |
| TRGHIGHFL | Y | 110 | 88 | 0.740 | 0.487 | 1.124 | 0.158 |
| TRGHIGHFL | N | 110 | 62 | 0.599 | 0.364 | 0.985 | 0.043 |

Subgroup-specific hazard ratios for overall survival are summarized across prespecified baseline categories. This table supports exploratory evaluation of consistency of treatment effect across clinically relevant strata.

**Supplementary Table S6.**
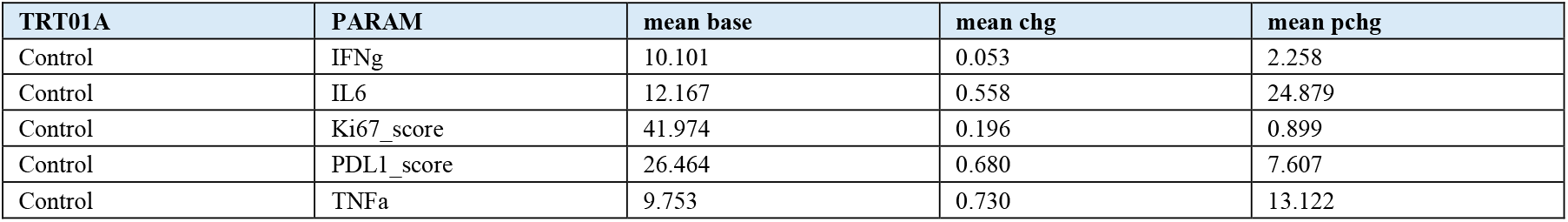
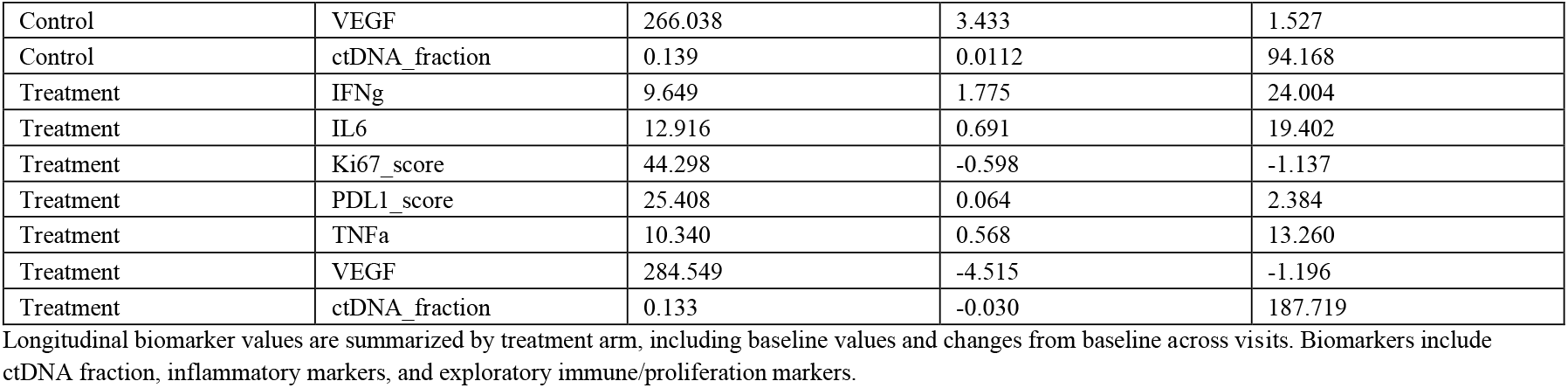
Biomarker change over time by treatment arm.

**Supplementary Table S7.**
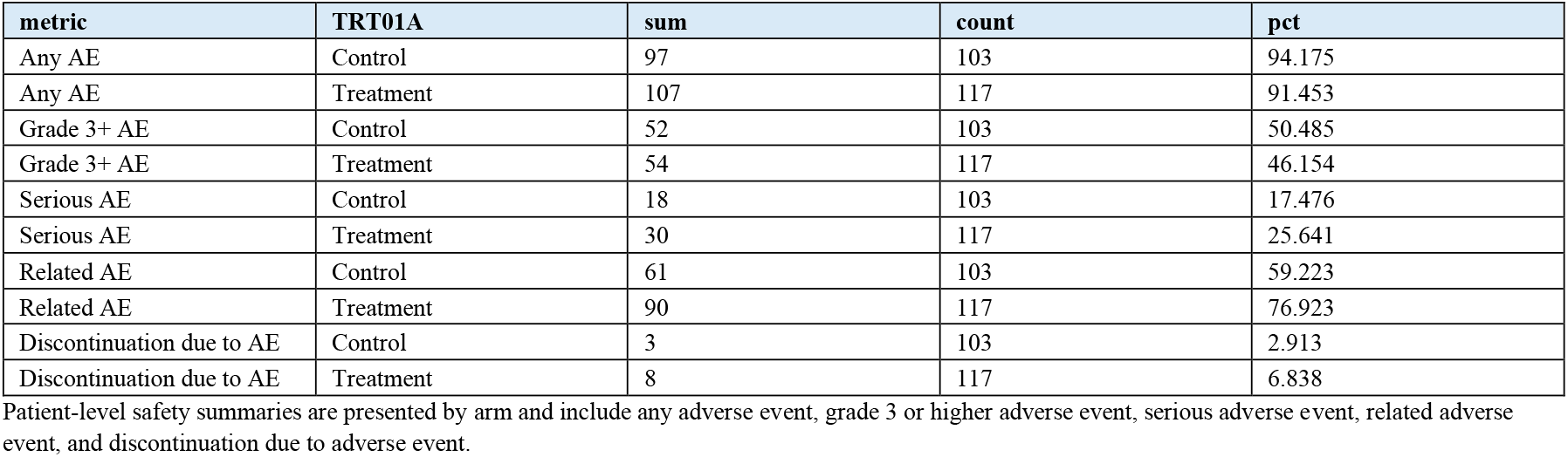
Adverse event summary by treatment arm.

**Supplementary Table S8.** Key laboratory parameters over time by treatment arm.

| TRT01A | PARAM | mean base | mean chg | mean pchg |
| --- | --- | --- | --- | --- |
| Control | ALT | 28.4072 | 0.331 | 6.223 |
| Control | AST | 24.695 | 0.401 | 10.659 |
| Control | Albumin | 3.929 | -0.054 | -1.364 |
| Control | CRP | 12.014 | 0.373 | 40.500 |
| Control | Creatinine | 0.969 | 0.002 | 0.646 |
| Control | Hemoglobin | 12.377 | -0.305 | -2.371 |
| Control | LDH | 249.924 | 0.630 | 0.599 |
| Control | Neutrophils | 4.258 | 0.010 | 1.074 |
| Control | Platelets | 257.912 | 0.611 | 0.563 |
| Treatment | ALT | 30.799 | 0.343 | 8.185 |
| Treatment | AST | 24.539 | -0.231 | 8.042 |
| Treatment | Albumin | 3.957 | -0.073 | -1.858 |
| Treatment | CRP | 12.328 | -0.357 | 8.650 |
| Treatment | Creatinine | 0.995 | 0.003 | 1.054 |
| Treatment | Hemoglobin | 12.376 | -0.341 | -2.640 |
| Treatment | LDH | 270.530 | -6.212 | -1.835 |
| Treatment | Neutrophils | 4.206 | -0.507 | -12.618 |
| Treatment | Platelets | 268.639 | -7.862 | -2.912 |

Longitudinal laboratory parameters are summarized by treatment arm. For each parameter, baseline values and post-baseline changes are shown to illustrate trends during follow-up.

## Notes

### Competing Interest Statement

The authors have declared no competing interest.

### Clinical Protocols

https://github.com/mpetalcorin/synthetic-oncology-clinical-trial-framework

### Funding Statement

This study did not receive any funding.

### Author Declarations

This work use ONLY simulated data openly available in the GitHub repository using this link: https://github.com/mpetalcorin/synthetic-oncology-clinical-trial-framework.

